# Clinical features and outcomes of 2019 novel coronavirus-infected patients with high plasma BNP levels

**DOI:** 10.1101/2020.03.31.20047142

**Authors:** Youbin Liu, Dehui Liu, Huafeng Song, Chunlin Chen, Mingfang Lv, Xing Pei, Zhongwei Hu, Zhihui Qin, Jinglong Li

**Author notes:** Corresponding author: Youbin Liu, MD; Department of Cardiology, Guangzhou Eighth People’s Hospital, Guangzhou medical university,8 huaying road, Baiyun district, Guangzhou city, Guangdong province (510000), China, Jinglong Li;Department of Cardiology, Guangzhou Eighth People’s Hospital, Guangzhou medical university,8 huaying road, Baiyun district, Guangzhou city, Guangdong province (510000), China. These authors are equal to the work□ (Youbin Liu, Dehui Liu, Huafeng Song, Chunlin Chen, Jinglong Li). **Funding/Support:** None.

## Abstract

**Aims:** To explore clinical features and outcome of 2019 novel coronavirus(2019-nCoV)-infected patients with high BNP levels

**Methods and results:** Data were collected from patients’ medical records, and we defined high BNP according to the plasma BNP was above > 100 pg/mL. In total,34 patients with corona virus disease 2019(COVID-19)were included in the analysis. Ten patients had high plasma BNP level. The median age for these patients was 60.5 years(interquartile range, 40-80y), and 6/10 (60%) were men. Underlying comorbidities in some patients were coronary heart disease (n=2, 20%), hypertesion (n=3,30%), heart failure (n=1,10%)and diabetes (n=2, 20%). Six (60%) patients had a history of Wuhan exposure. The most common symptoms at illness onset in patients were fever (n=7, 70%), cough (n=3, 30%), headache or fatigue(n=4,40%). These patients had higher aspartate aminotransferase(AST), troponin I, C reactive protein and lower hemoglobin, and platelet count,compared with patients with normal BNP, respectively. Compared with patients with normal BNP, patients with high BNP were more likely to develop severe pneumonia, and receive tracheal cannula, invasive mechanical ventilation, continuous renal replacement therapy, extracorporeal membrane oxygenation, and be admitted to the intensive care unit. One patient with high BNP died during the study.

**Conclusion:** High BNP is a common condition among patients infected with 2019-nCoV. Patients with high BNP showed poor clinical outcomes

## INTRODUCTION

The 2019 novel coronavirus (2019-nCoV), a new fatal virus that emerged at the end of 2019, is a growing public health concern worldwide (1). The findings from previous studies show that some infected patients had abnormal laboratory test results, including blood cell counts, BNPand brain natriuretic peptide(BNP) and so on (2) (3). However, as a new coronavirus, we still know little about whether 2019-nCoV is more likely to be harmful for these patients with with high plasma BNP levels and the role of BNP in corona virus disease 2019(COVID-19). More detailed investigations of the relationship between BNP and the clinical outcomes of people infected with 2019-nCoV are urgly needed.

Brain natriuretic peptide (BNP), a member of a family of natriuretic peptides, was first idendified in 1988 and discovered to be present in high concentrations in cardiac tissues,, especially the ventricles (4). Initial studies showed that BNP levels were strongl related to impaired left ventricular (LV) function (5). In recent years, BNP, as a valuable clinical biochemical marker, has been widely used in the diagnosis, prognosis and therapeutic effect evaluation of cardiovascular diseases such as acute coronary syndrome, right ventricular dysfunction,pulmonary disease, diastolic dysfunction (6).However, plasma BNP levels are affected by many factors. BNP is not only regulated by myocardial extension, but also affected by factors such as tachycardia, epinephrine, thyroxine, vasoactive peptide and infection and so on (7). 2019-nCoV, as a new virus, although it mainly damages lung tissue, some studies have found that it also has a destructive effect on the heart. A recent study indicate N-Terminal pro-brain natriuretic peptide(NT-proBNP) has a prognostic value in severe covid-19 patients (8).However, The role of BNP in COVID-19 patient is still unknown..

This paper provides an overview of the clinical features of 2019-nCoV-infected patients with high plasma BNP levels to provide insight into the prevention and treatment for these patients.

## METHODS

In total,34 patients with corona virus disease 2019(COVID-19)were included in analysis. Patients were admitted to Guangzhou eighth people’s hospital from January 20, 2020, to February 24, 2020. Throat swab specimens were gathered from all patients after admission, and Real-Time polymerase chain reaction were performed to detect 2019-nCoV ribonucleic acid. All patients with COVID-19 were diagnosed based on the World Health Organization interim guidelines(9). Unless otherwise specified, all values are the first data after admission, and if the index was measured more than twice, we chose the highest value for analysis. High BNP level was diagnosed if the plasma BNP were above the 99th percentile of the upper reference limit (> 100 pg/mL)) using the tridge BNP test (Beckman Coulter Inc., Brea, CA, USA). Pneumonia severity was defined according to the international guidelines for community-acquired pneumonia(10). The epidemiological, laboratory, clinical and outcome data are derived from the patient’s electronic medical records. the ethics commissions of the Guangzhou Eighth people’s hospital has approved this study, with a waiver of informed consent.

Continuous variables were expressed as mean ± standard deviation for normally distributed data or as median (interquartile range,IQR) for skewed distributions. Frequency data were presented as proportions. Student’s t test or the Mann–Whitney U test were performed for continuous variables when appropriate, whereas differences in categorical variables were assessed using the Chi-square test or Fisher’s exact test. SPSS 25.0 (IBM Corp. Armonk, NY, USA) were used to perform All analyses and a two-tailed p-value < 0.05 was considered statistically significant.

## RESULTS

### 1. Epidemiological features of 2019-nCoV-infected patients with high BNP levels

A total of 34 patients were included and divided into two groups (high BNP group and normal group) in the final analysis. Ten patients had high plasma BNP level (>100pg/mL). The median age for these patients was 60.5 years(interquartile range, 40-80y), and 6/10 (60%) were men. Underlying comorbidities in some patients were coronary heart disease (n=2, 20%), hypertesion (n=3,30%), heart failure (n=1,10%)and diabetes (n=2, 20%). Six (60%) patients had a history of wuhan exposure. The epidemiological characteristics of the study participants are presented in Table 1.

**Table 1.** The epidemiological features of 2019-nCoV-infected patients with high BNP levels.

| Variables | BNP>100<br>(N=10) | BNP≤100<br>(N=24) | P |
| --- | --- | --- | --- |
| Age (Y) | 60.5(40-80) | 38(33-52) | 0.02* |
| Male, No(%) | 6 ( 60 ) | 12 ( 50 ) | 0.72 |
| Exposure history in Wuhan, No(%) | 6 ( 60 ) | 6(25) | 0.11 |
| Diabetes, No(%) | 2 ( 20 ) | 2(8.3) | 0.56 |
| Coronary heart disease, No(%) | 2(20) | 3(12.5) | 0.62 |
| Heart failure, No(%) | 1(10) | 0(0) | 0.29 |
| Arrhythmia, No(%) | 1(10) | 0(0) | 0.29 |

### 2. Clinical features and laboratory findings of 2019-nCoV-infected patients with high BNP levels

The most common symptoms at illness onset in patients were fever (n=7, 70%), cough (n=3, 30%), headache or fatigue (n=4,40%). These patients had higher aspartate amino transferase(AST),troponin I,C reactive protein and lower hemoglobin,and platelet count,compared with patients with normal BNP, respectively. The clinical features and selected laboratory findings of the study participants are presented in Table 2.

**Table 2.** The clinical features and selected laboratory findings of 2019-nCoV-infected patients with high BNP levels.

| Variables | Normal<br>range | BNP>100<br>(N=10) | BNP≤100<br>(N=24) | *P* |
| --- | --- | --- | --- | --- |
| Cough, No(%) | - | 3(30) | 15(62.5) | 0.13 |
| Fever, No(%) | - | 7(70) | 11(45.8) | 0.27 |
| Sore throat ,No(%) | - | 1 ( 10 ) | 6(25) | 0.64 |
| Headache or Fatigue, No(%) | - | 4(40) | 3(12.5) | 0.16 |
| Dyspnea, No(%) | - | 2(20) | 5( 20.8 ) | 1.0 |
| Chest pain , No(%) | - | 0(0) | 0(0) | 1.0 |
| Palpitation, No(%) | - | 0(0) | 1(4.2) | 1.0 |
| Heart rate ( bpm ) | 55-95 | 83(72-98) | 87.5(80-97) | 0.49 |
| Temperature (°C) | 36.3-37.3 | 36.9(36.5-38.3) | 36.5(36-37) | 0.09 |
| Systolic pressure(mmHg) | 90-139 | 134(121-145) | 127(110-147) | 0.38 |
| White blood cell count (10E9/L) | 4-10 | 5.6(5.0-10.2) | 5.3(3.8-6.4) | 0.45 |
| Neutrophil count (10E9/L) | 1.8-6.3 | 3.7(3.2-7.0) | 3.1(2.1-4.3) | 0.27 |
| Lymphocytes count (10E9/L) | 1.1-3.2 | 1.0(0.7-2.2) | 1.3(1.1-1.9) | 0.17 |
| Hemoglobin, g/L | 113-151 | 115.3±27.5 | 132.4±18 | 0.04* |
| Platelets count (10E9/L) | 100-300 | 155.6±57.2 | 226.38±47.1 | 0.001* |
| C reactive protein (>10mg/L) , No(%) | <10 | 9(90) | 5( 20.8) | 0.00* |
| Troponin I, ( ug/L) | <0.03 | 0.08(0.02-0.29) | 0.005(0.001-0.01) | 0.00* |
| Creatinine, μmol/L | 59~104 | 63.1(43.6-137.8) | 60.5(51.4-77.0) | 0.809 |
| Brain natriuretic peptide (pg/mL) | <100 | 245.5(142.5-371.8) | 18(9.8-36.3) | 0.00* |
| Aspartate aminotransferase (U/L) | 13-35 | 24.7(18.95-37.6) | 17.6(13.4-19.95) | 0.02* |
| Alanine aminotransferase (U/L) | 7-40 | 16.3(9.8-24.8) | 16.6(12.8-24.5) | 0.59 |
| Myoglobin (ug/L ) | 17.4-105.7 | 32.8(15.8-156.2) | 15.1(11-20.9) | 0.001* |
| Creatine kinase (U/L) | 50-310 | 48(34.8-83.8) | 56.2(42.3-78.3) | 0.86 |
| D-dimer(mg/L) | <1000 | 1765(667.5-6085) | 990(660-1280) | 0.15 |
| Bilateral pneumonia No(%) | -- | 9(90) | 14(58.3) | 0.11 |

**Table 3.**
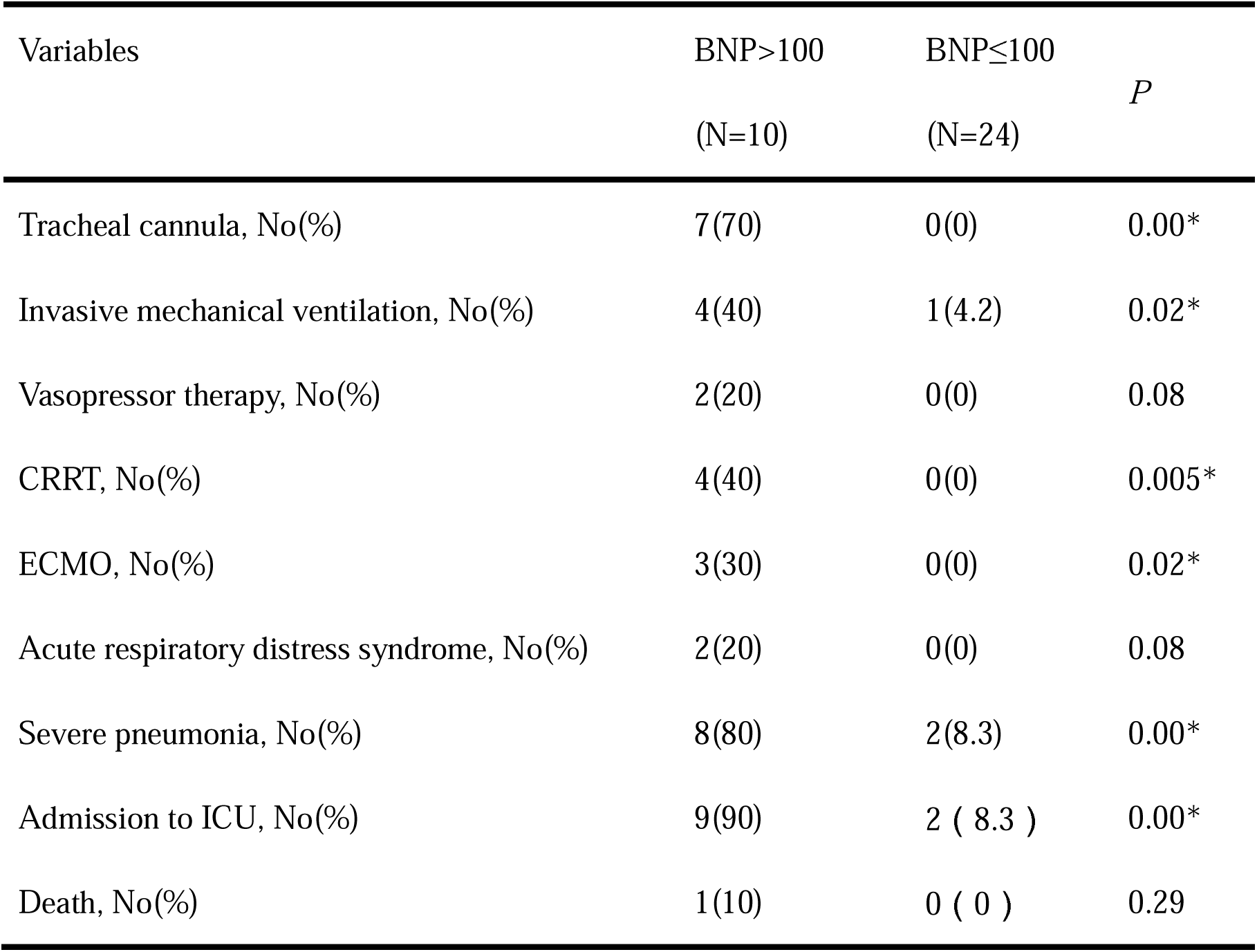
Treatments and outcomes of 2019-nCoV-infected patients with high BNP levels.

| Variables | BNP>100<br>(N=10) | BNP≤100<br>(N=24) | *P* |
| --- | --- | --- | --- |
| Tracheal cannula, No(%) | 7(70) | 0(0) | 0.00* |
| Invasive mechanical ventilation, No(%) | 4(40) | 1(4.2) | 0.02* |
| Vasopressor therapy, No(%) | 2(20) | 0(0) | 0.08 |
| CRRT, No(%) | 4(40) | 0(0) | 0.005* |
| ECMO, No(%) | 3(30) | 0(0) | 0.02* |
| Acute respiratory distress syndrome, No(%) | 2(20) | 0(0) | 0.08 |
| Severe pneumonia, No(%) | 8(80) | 2(8.3) | 0.00* |
| Admission to ICU, No(%) | 9(90) | 2 ( 8.3 ) | 0.00* |
| Death, No(%) | 1(10) | 0 ( 0 ) | 0.29 |

### Treatments and outcomes of 2019-nCoV-infected patients with high BNP levels

Compared with patients with normal BNP, patients with high BNP were more likely to develop severe pneumonia (80% vs 8.3%), and receive tracheal cannula(70% vs 0%), invasive mechanical ventilation (40% vs 4.2%), continuous renal replacement therapy(40% vs 0%), extracorporeal membrane oxygenation(30% vs 0%), and be admitted to the intensive care unit (90% vs 8.3%). One patient with high BNP died during the study.The epidemiological characteristics and outcomes of the study participants are presented in Table.

## DISCUSSION

To the best of our knowledge, this is the first study systematically exploring clinical features and outcomes of 2019 novel coronavirus-infected patients with high BNP(not NT-proBNP) levels. Our results showed that 2019-nCoV infected patients with high plasma BNP levels had worse clinical outcomes compared with patients with normal plasma BNP levels.

BNP and NT-proBNP, peptides produced by cardiomyocytes are widely used to guide in diagnosis, prognosis and treatment of heart failure (11). It is well known that the level of BNP in plasma is affected by many factors, such as inflammation and stress reaction and so on (12).Therefore, it is very common for patients with other disease are often accompanied with high plasma BNP level. Some studies have shown patients with COVID-19 often had abnormal BNP/NT-proBNP in plasma (2) (3).Howerver,by now,there is no detailed investigation on clinical features and outcomes of 2019 novel coronavirus-infected patients with high BNP levels.This study provides information on the epidemiology and outcomes of 2019-nCoV-infected patients with high plasma BNP levels. Most of patients with high BNP levels in our study were usually older and often had pre-existing heart disease. High BNP level following 2019-nCoV infection is associated with poor patient outcomes. These patients were more likely receive mechanical ventilation, tracheal cannula, continuous renal replacement therapy, extracorporeal membrane oxygenation and be admitted to the intensive care unit.

However, the reason why the outcomes of patients with high BNP were worse is unclear. Inflammation and stress can stimulate BNP production and secretion from cardiomyocytes(12).The level of BNP in plasma may reflect the severity of inflammation and stress. This may partly explain why patients with high plasma BNP levels had a bad outcomes. This study is limited by a relatively small number of samples from patients with high BNP.These data contribute information to understanding clinical manifestations and outcomes of 2019-nCoV infected patients.

## Data Availability

All data will be available when be requested

## Acknowledgements

The authors had full access to all of the data in the study and take responsibility for the integrity of the data and the accuracy of the data analysis. All authors have no conflict of interest to declare.

